# Medical discrimination and the selective erosion of institutional health trust: evidence from the Health Information National Trends Survey 6 and 7

**DOI:** 10.64898/2026.06.06.26355057

**Authors:** Adam Park, Luke Yin, Andrew Wong, Cooper Lee, Yoo Choi

## Abstract

Medical discrimination may alter how patients relate to health information sources following adverse care encounters. We examined whether discrimination experience is associated with selective erosion of institutional health trust and with compensatory digital health engagement, using nationally representative data from the Health Information National Trends Survey (HINTS) 6 (2022; n=6,252) and HINTS 7 (2024; n=7,278). Survey-weighted modified Poisson regression estimated prevalence ratios (PRs) for binary high-trust outcomes, and survey-weighted ordinary least squares estimated coefficients for continuous outcomes; jackknife replicate weights (50 replicates) provided variance estimates. Discrimination was associated with substantially lower probability of high trust in the healthcare system (PR=0.39; 95% CI 0.30–0.52) and physicians (PR=0.85; 95% CI 0.77–0.94), with no significant association for trust in scientists, government, family, or religious organisations. The clinical-institutional pattern replicated in HINTS 6, which additionally showed reduced trust in scientists for race/ethnicity-based discrimination. Contrary to a disengagement hypothesis, discrimination-exposed adults showed higher probability of online health information seeking (PR=1.06), health app use (PR=1.11), and online provider messaging (PR=1.13); these associations persisted after adjustment for trust in physicians. Discrimination was independently associated with lower health self-efficacy (b=−0.271). Medical discrimination selectively erodes trust in clinical institutions while leaving broader epistemic trust largely intact. Despite this, discrimination-exposed patients engage more actively with digital health channels, consistent with compensatory reorientation toward non-clinical information sources. These findings describe engaged but institutionally alienated patients, with implications for restoring clinical trust and for equity-centred digital health design.

## Introduction

Discrimination in medical care is not an exceptional experience for marginalised Americans. National surveys consistently document that racial and ethnic minority patients, lower-income adults, and those with stigmatised health conditions report differential treatment, inadequate communication, and reduced care quality when seeking healthcare [1–4]. These encounters carry consequences that extend beyond the immediate visit: they reshape the cognitive and affective frameworks patients bring to subsequent interactions, including how much they trust the systems and sources from which they seek health information.

The relationship between discrimination experience and trust in healthcare providers and institutions has received substantial scholarly attention [5–10]. Prior work documents associations between perceived discrimination and reduced likelihood of engaging preventive care [11, 12], avoiding clinical interactions [13], and lower satisfaction with care quality [14]. What remains less well understood is the architecture of trust erosion: whether discrimination produces broad distrust across all health information sources, or whether its effects are selective, targeting only the institutional contexts where the discrimination occurred.

This distinction matters considerably. If discrimination erodes trust specifically in physicians and the healthcare system while leaving trust in scientists, public health authorities, and digital health platforms intact, then discrimination-exposed patients may rationally redistribute their health information seeking toward non-clinical channels. This would constitute compensatory channel-switching: a strategic adaptation in which institutional trust erosion redirects engagement toward perceived alternatives [15, 16], rather than producing disengagement from health information generally.

The Health Information National Trends Survey (HINTS), conducted by the National Cancer Institute, captures detailed information about both discrimination experience and digital health engagement across its two most recent waves (HINTS 6, 2022; HINTS 7, 2024) [17–19]. With a combined analytic sample of more than 13,000 adults, HINTS offers sufficient statistical power to model the relationships among discrimination, source-specific trust, digital health engagement, and patient health outcomes. We pursued three objectives: to characterise the trust-specificity of discrimination’s effects across six source domains; to test whether discrimination-exposed adults show different patterns of digital health engagement; and to examine whether discrimination is independently associated with patients’ confidence in managing their own health.

## Materials and methods

### Ethics statement

This study involved secondary analysis of de-identified, publicly available data from the Health Information National Trends Survey and did not constitute human-subjects research requiring institutional review board approval.

### Study design and data sources

We used publicly available data from HINTS 6 (2022; n=6,252) and HINTS 7 (2024; n=7,278), which are freely available from the National Cancer Institute (https://hints.cancer.gov). HINTS is a nationally representative cross-sectional survey of non-institutionalised U.S. adults. Both waves used address-based sampling with web and mail response options and calibration weighting to produce nationally representative estimates [17, 18]. We analysed the two waves separately, treating HINTS 7 as the primary sample and HINTS 6 as a historical comparison.

### Discrimination exposure

In HINTS 6, discrimination was measured by the item: “Have you ever been treated unfairly or been discriminated against when getting medical care because of your race or ethnicity?” (Yes/No; DiscriminatedMedCare). In HINTS 7, the question was broadened: “Have you ever experienced prejudice or been discriminated against when getting medical care?” (Yes/No; DiscriminatedMedCare2). This wording change removes the race/ethnicity qualifier, capturing discrimination on any basis. The broadened measure in HINTS 7 serves as the primary exposure; HINTS 6 provides a comparison using the narrower racial/ethnic discrimination construct.

### Trust outcomes

Trust in health information sources was assessed using items asking respondents how much they would trust information about cancer from physicians/doctors, scientists, government health agencies, the healthcare system, family and friends, and religious organisations (1=Not at all to 4=A lot). Each item was dichotomised as high trust (A lot) versus lower trust (all other responses). Endorsement of high trust ranged from common (physicians, approximately 71%) to relatively rare (religious organisations, approximately 4%). We estimated prevalence ratios using modified Poisson regression with a log link, which provides directly interpretable prevalence ratios across the full prevalence range and, for the more commonly endorsed outcomes, avoids the overstatement of associations produced by odds ratios [20].

### Digital health engagement and health self-efficacy

Three binary digital health engagement outcomes were examined in HINTS 7: using the internet for health information in the past 12 months, using health and wellness apps, and sending messages to a provider through an online portal. Health self-efficacy was measured by the item “Overall, how confident are you about your ability to take good care of your health?” (1=Not confident at all to 5=Completely confident).

### Covariates

All models adjusted for age (continuous, centred at 50 years), sex (female=1), race/ethnicity (Non-Hispanic White reference; Non-Hispanic Black, Hispanic, Non-Hispanic Asian, Non-Hispanic Other indicators), education (4-level ordinal), household income (ordinal; 5 categories in HINTS 6 and 6 categories in HINTS 7), and insurance status (insured=1). Missing income data (approximately 11–12% per wave) were replaced with the median; sensitivity analyses with complete cases confirmed result stability.

### Statistical analysis

All analyses incorporated HINTS person-level weights and 50 jackknife replicate weights. Variance was estimated using the delete-1 jackknife formula, 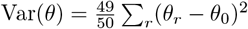 [21]. Survey-weighted modified Poisson regression (generalised linear model with log link) estimated PRs for binary outcomes [20]. Survey-weighted ordinary least squares estimated regression coefficients for continuous outcomes. The self-efficacy model included trust in the healthcare system, trust in physicians, two discrimination *×* trust interaction terms, digital health engagement variables, and all demographic and socioeconomic covariates. All analyses were conducted in Python 3.12 using statsmodels and custom jackknife weighting functions. Statistical significance was defined as a two-sided *α* of 0.05. Race and ethnicity were modelled using respondents’ self-reported classification from the HINTS five-category race/ethnicity variable, with Non-Hispanic White as the reference; socioeconomic confounding was addressed by adjusting for education, household income, and insurance status. Given the focused, theory-driven hypotheses and the replication of the primary trust associations across two independent survey waves, we did not apply a formal correction for multiple comparisons; associations for secondary outcomes should be interpreted as exploratory. Full regression results for every model, including all covariates, standard errors, and confidence intervals, are provided in S1 File. The study is reported in accordance with the STROBE guidelines for observational studies (S2 File).

## Results

### Sample characteristics

Table 1 presents weighted descriptive statistics for both waves. Samples were broadly similar in demographic composition: approximately 60% Non-Hispanic White, 11% Non-Hispanic Black, 17% Hispanic, and 6% Non-Hispanic Asian; mean age approximately 49 years. Weighted discrimination prevalence was 7.3% (95% CI 6.3–8.3%) in HINTS 6 and 16.1% (95% CI 14.3–17.9%) in HINTS 7, consistent with the broadened question wording. Trust in physicians was the most commonly endorsed high-trust response in both waves (approximately 71%), followed by trust in scientists (approximately 54%), while trust in the healthcare system (approximately 35%) was notably lower.

**Table 1.** Weighted descriptive statistics, HINTS 6 (2022) and HINTS 7 (2024).

| Characteristic | HINTS 6 (2022)<br>n=6,252 | HINTS 7 (2024)<br>n=7,278 |
| --- | --- | --- |
| Discrimination prevalence, % (95% CI) | 7.3 (6.3–8.3) | 16.1 (14.3–17.9) |
| *Trust in information sources (% high trust)* |  |  |
| Physicians/doctors | 72.2 | 71.1 |
| Scientists | 54.8 | 54.3 |
| Government agencies | 27.8 | 26.8 |
| Healthcare system | 36.0 | 33.8 |
| Family and friends | 7.9 | 6.3 |
| Health self-efficacy, mean (SD) | 3.87 (0.92) | 3.83 (0.87) |
| Health literacy, mean (SD) | 3.39 (0.84) | 3.51 (0.78) |
| *Digital health behaviours* |  |  |
| Online health information seeking, % | 84.7 | 85.2 |
| Health app use, % | 56.6 | 58.3 |
| *Demographics* |  |  |
| Age, mean years (SD) | 48.7 (18.0) | 49.0 (18.0) |
| Female, % | 50.8 | 48.8 |
| Non-Hispanic White, % | 61.2 | 60.6 |
| Non-Hispanic Black, % | 11.0 | 11.1 |
| Hispanic, % | 17.0 | 17.5 |
| Non-Hispanic Asian, % | 5.7 | 5.6 |
| College graduate or more, % | 32.6 | 33.9 |
| Insured, % | 89.3 | 91.4 |

All estimates are survey-weighted. The discrimination prevalence 95% CI was estimated using jackknife replicate weights. Trust percentages reflect the weighted proportion reporting “A lot” of trust. Health literacy reflects a single-item measure of confidence completing medical forms (scale 1–4); health self-efficacy is scaled 1–5. HINTS = Health Information National Trends Survey.

### Medical discrimination and institutional trust

Table 2 presents HINTS 7 results. Discrimination was associated with substantially lower probability of high trust in the healthcare system (PR=0.39; 95% CI 0.30–0.52; p<0.001) and lower probability of high trust in physicians (PR=0.85; 95% CI 0.77–0.94; p=0.001). Discrimination-exposed adults were 61% less likely to report high trust in the healthcare system and 15% less likely to report high trust in physicians relative to non-exposed adults. Discrimination was not significantly associated with trust in scientists, government agencies, family and friends, or religious organisations (all p>0.20), indicating selective rather than diffuse erosion of trust. Fig 1 illustrates these associations across both waves.

**Table 2.**
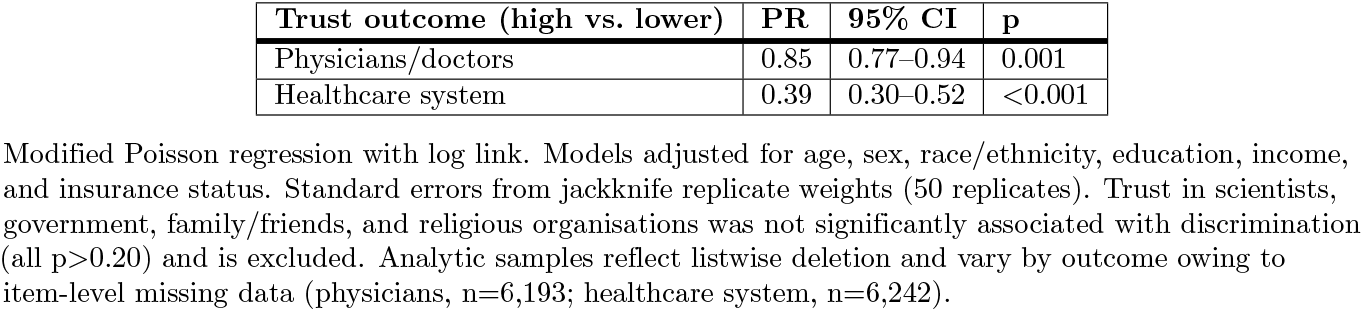
Medical discrimination and institutional trust: HINTS 7 (2024) primary analysis (n=6,193–6,242).

**Fig 1.**
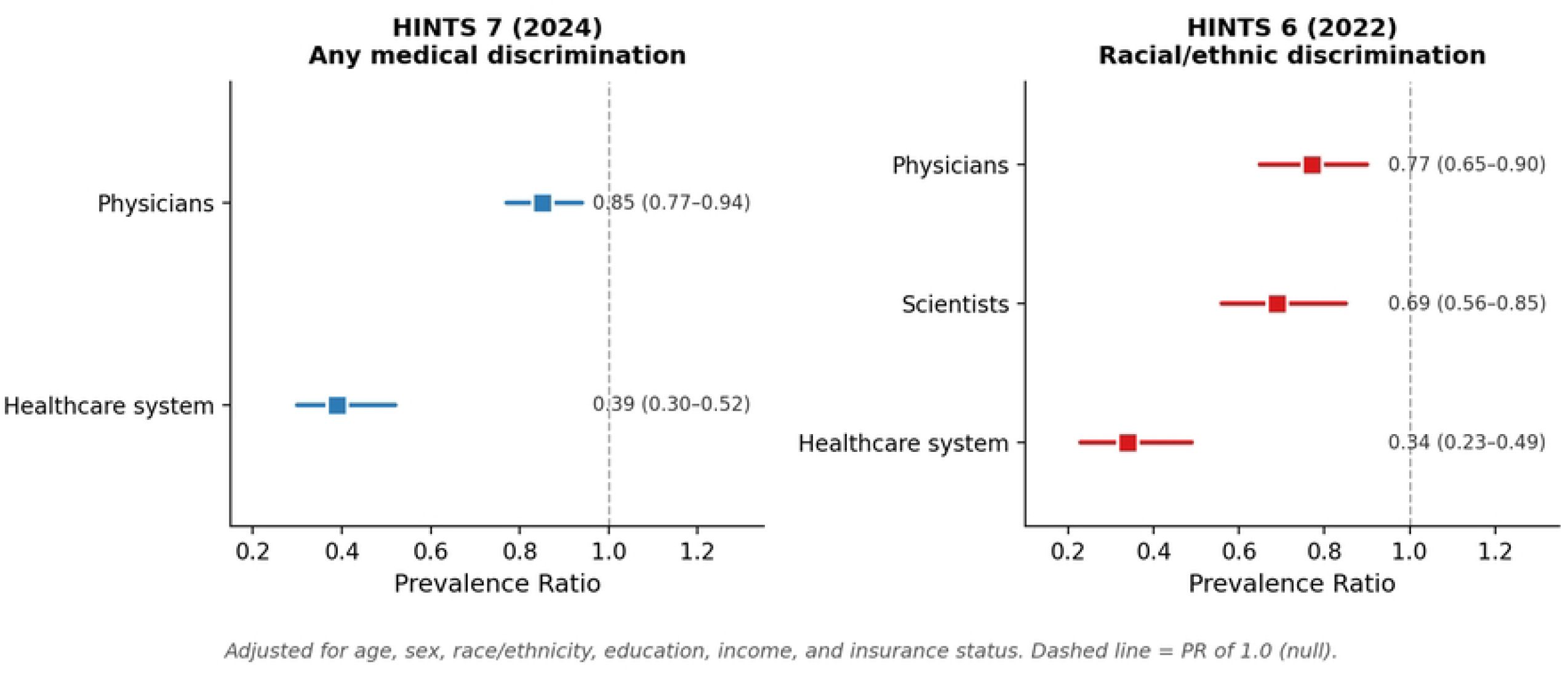
Medical discrimination and institutional trust: forest plots from HINTS 6 and HINTS 7. Squares represent prevalence ratios; horizontal lines represent 95% confidence intervals. Values are shown as PR (95% CI). The dashed line marks a prevalence ratio of 1.0 (null). Only statistically significant trust outcomes are displayed. Models were adjusted for age, sex, race/ethnicity, education, income, and insurance status.

Table 3 presents HINTS 6 results, in which discrimination was measured specifically as race/ethnicity-based. The clinical-institutional pattern replicated, with the healthcare system (PR=0.34; 95% CI 0.23–0.49; p<0.001) and physicians (PR=0.77; 95% CI 0.65–0.90; p=0.001) again showing the strongest effects. In HINTS 6, a significant association with scientist trust also emerged (PR=0.69; 95% CI 0.56–0.85; p=0.001), which was not present in HINTS 7. This may reflect that race/ethnicity-based discrimination carries a broader epistemic dimension implicating scientific institutions perceived as complicit in structural racism [22, 23].

**Table 3.** Racial/ethnic medical discrimination and trust: HINTS 6 (2022) historical comparison (n=5,287–5,430).

| Trust outcome (high vs. lower) | PR | 95% CI | p |
| --- | --- | --- | --- |
| Physicians/doctors | 0.77 | 0.65–0.90 | 0.001 |
| Scientists | 0.69 | 0.56–0.85 | <0.001 |
| Healthcare system | 0.34 | 0.23–0.49 | <0.001 |

### Discrimination and digital health engagement

Table 4 and Fig 2 present results from models predicting digital health engagement. Contrary to a disengagement hypothesis, discrimination-exposed adults showed significantly higher probability of online health information seeking (PR=1.06; 95% CI 1.02–1.10; p=0.001), health app use (PR=1.11; 95% CI 1.02–1.20; p=0.012), and online provider messaging (PR=1.13; 95% CI 1.05–1.21; p=0.001). These associations persisted after adjusting for trust in physicians, indicating that elevated digital engagement is not simply a proxy for lower physician trust.

**Table 4.** Medical discrimination and digital health engagement: HINTS 7 (2024) (n=5,952–5,995).

| Digital health outcome | PR | 95% CI | p |
| --- | --- | --- | --- |
| Online health information seeking | 1.06 | 1.02–1.10 | 0.001 |
| Health app use | 1.11 | 1.02–1.20 | 0.012 |
| Online provider messaging | 1.13 | 1.05–1.21 | 0.001 |

Modified Poisson regression. Models adjusted for trust in physicians (continuous), age, sex, race/ethnicity, education, income, and insurance status. Analytic samples reflect listwise deletion and vary by outcome (online information seeking, n=5,957; health app use, n=5,995; online provider messaging, n=5,952).

**Fig 2.**
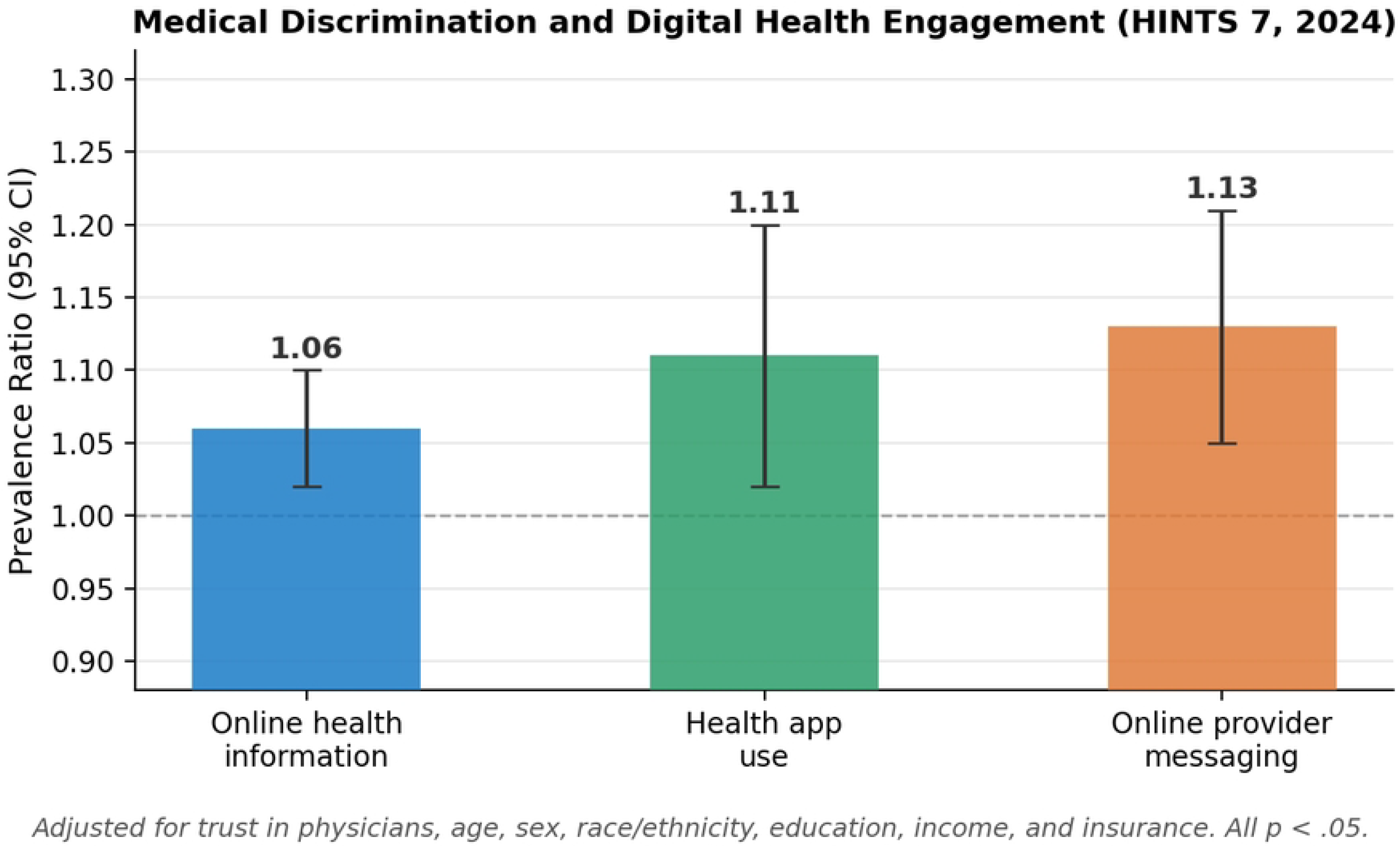
Medical discrimination and digital health engagement (HINTS 7, 2024). Bars represent prevalence ratios; error bars represent 95% confidence intervals. The dashed line marks a prevalence ratio of 1.0. Models were adjusted for trust in physicians, age, sex, race/ethnicity, education, income, and insurance status. All associations p<0.05.

### Health self-efficacy outcome model

Table 5 and Fig 3 present results from the self-efficacy model. Medical discrimination was independently associated with lower health self-efficacy (b=−0.271; 95% CI −0.402 to −0.140; p<0.001). Trust in the healthcare system was positively associated with self-efficacy (b=0.182; 95% CI 0.107–0.258; p<0.001). Health app use was positively associated with self-efficacy (b=0.104; 95% CI 0.022–0.186; p=0.013), while online health information seeking was negatively associated (b=−0.195; 95% CI −0.299 to −0.091; p<0.001). Income was also independently associated with higher self-efficacy (b=0.060; p<0.001).

**Table 5.** Health self-efficacy outcome model: HINTS 7 (2024), survey-weighted OLS (n=5,796).

| Predictor | b | 95% CI | p |
| --- | --- | --- | --- |
| Medical discrimination | −0.271 | −0.402 to −0.140 | <0.001 |
| Trust in healthcare system | 0.182 | 0.107–0.258 | <0.001 |
| Health app use | 0.104 | 0.022–0.186 | 0.013 |
| Online health information seeking | −0.195 | −0.299 to −0.091 | <0.001 |
| Age (per year above 50) | −0.003 | −0.005 to −0.000 | 0.029 |
| Income | 0.060 | 0.034–0.086 | <0.001 |

Outcome: health self-efficacy (1=Not at all confident to 5=Completely confident). The model also included two discrimination × trust interaction terms (discrimination × trust in physicians and discrimination × trust in healthcare system), both non-significant (p>0.34). Non-significant predictors (sex, insurance status, trust in physicians, education, and the two interaction terms) are omitted from the table. Race/ethnicity indicators and age were included in the model but are not all shown. The analytic sample comprised 5,796 respondents with complete data on all model variables.

**Fig 3.**
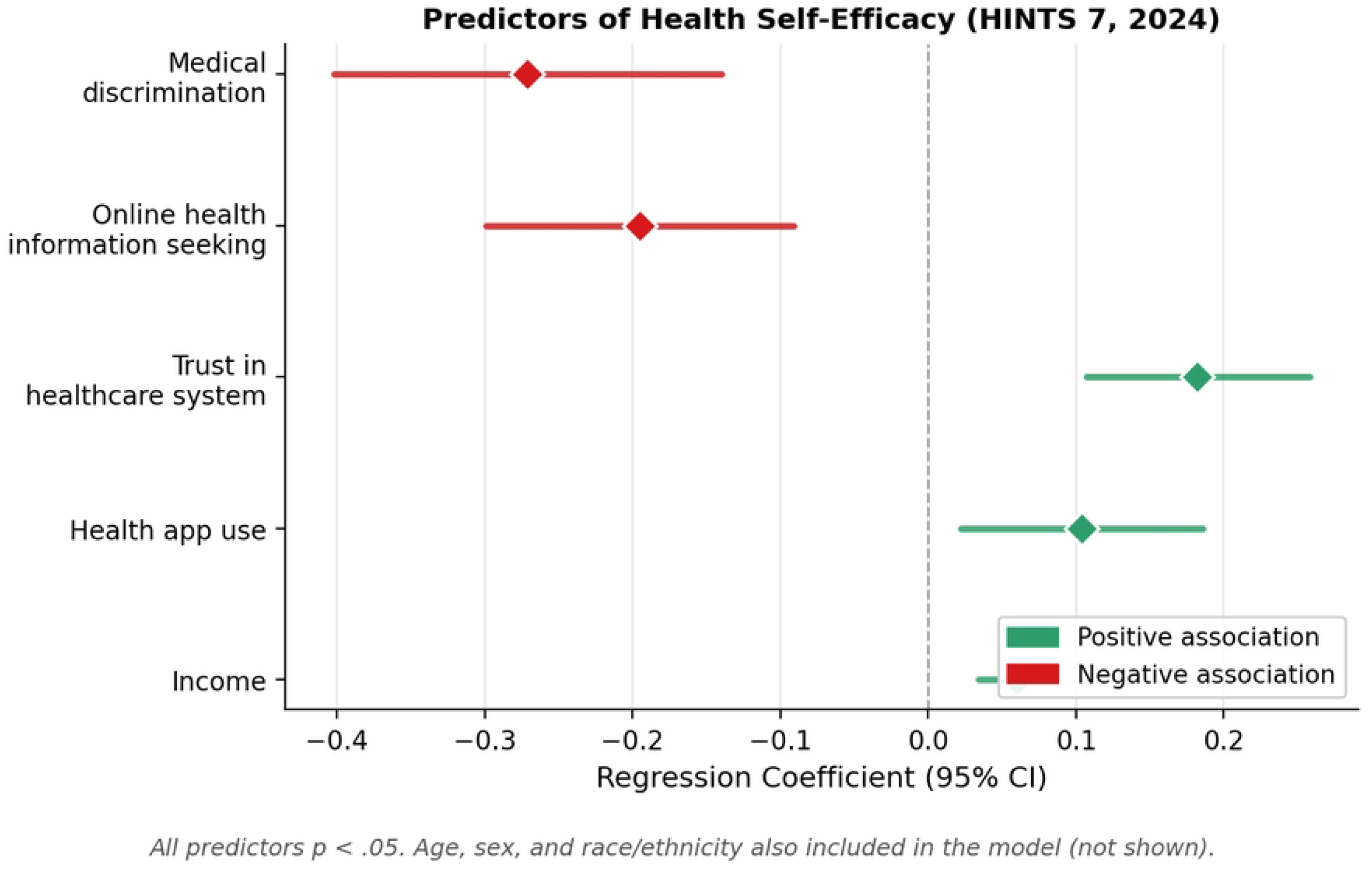
Significant predictors of health self-efficacy (HINTS 7, 2024). Diamonds represent regression coefficients (b); horizontal lines represent 95% confidence intervals. The dashed line marks a null effect. All predictors shown are p<0.05. Age, sex, and race/ethnicity were included in the model but are not displayed.

## Discussion

Using two consecutive waves of a nationally representative health information survey, this study documents that medical discrimination erodes trust in a selective, institutionally specific manner. Discrimination was strongly associated with lower trust in the healthcare system and physicians, the specific actors implicated in the discrimination experience, but was unrelated to trust in scientists, government agencies, family, or religious organisations. This clinical-institutional specificity was replicated across both HINTS waves, providing convergent evidence for the core finding.

The institutional specificity of trust erosion argues against a generalised alienation model in which discrimination produces broad epistemic distrust and instead supports what we term a bounded erosion model: discrimination experience damages trust in the precise institutional relationships where it occurs while leaving other trust relationships intact. This is consistent with social exchange theory, in which trust violations within a relationship alter assessments of that specific relationship without necessarily generalising to other domains [24]. Patients who have experienced discrimination in medical encounters may come to regard physicians and the healthcare system as unreliable while continuing to view scientists and public health information as credible.

The finding that race/ethnicity-based discrimination additionally erodes trust in scientists while broader medical care discrimination does not is substantively informative. Race/ethnicity-based discrimination may carry an epistemic dimension that extends beyond individual clinical encounters to implicate scientific institutions perceived as complicit in structural racism [22, 23]. Broader forms of medical discrimination may be more readily interpreted as individual provider failures without implicating the scientific establishment.

The compensatory digital engagement finding is operationally significant. Discrimination-exposed adults were more likely to use the internet for health information, health apps, and online provider messaging, and these associations persisted after adjusting for physician trust. The elevated digital engagement likely reflects an independent motivation to engage with health management through channels perceived as more controllable or less subject to discriminatory dynamics [15]. This aligns with accounts of patients using digital health information to prepare for clinical encounters, verify information received from providers, or compensate for perceived inadequacies in care. The inverse association between online health information seeking and self-efficacy in the outcome model may reflect reverse causality, with less-confident adults seeking online information more actively rather than online seeking lowering confidence.

The self-efficacy findings underscore the independent harm of discrimination exposure. Even after adjusting for trust levels and digital engagement, discrimination was associated with lower health self-efficacy. The pathway from discrimination to impaired self-efficacy likely involves mechanisms beyond trust, such as internalised stigma, reduced perceived competence following adversarial care encounters, or cumulative stress effects [25–27].

For health system leaders, these findings highlight the importance of addressing discrimination as a systemic concern. Trust in the healthcare system showed the largest discrimination-associated deficit: 61% lower high-trust probability in HINTS 7. Restoring this trust requires structural interventions, including diversity in the health workforce, transparent accountability mechanisms, and system-level signals that adverse care experiences will be taken seriously [28, 29]. For digital health designers and implementers, discrimination-exposed patients are not digitally disengaged. The positive association between health app use and self-efficacy suggests that digital tools can support patient activation even among those with eroded institutional trust. Ensuring these tools are designed with health equity principles is essential to translating motivated engagement into health benefit.

## Limitations

Several limitations warrant acknowledgment. The cross-sectional design precludes causal inference; reverse causality cannot be excluded. Discrimination measures are lifetime prevalence items that do not capture recency, frequency, or type, limiting dose-response characterisation. The question wording change between waves precludes direct prevalence comparison. Trust items reference cancer information specifically, which may introduce construct specificity relative to general health information trust. Future studies using continuous trust measures and prospective designs would strengthen the causal evidence base.

## Conclusion

Medical discrimination erodes trust in healthcare institutions with remarkable selectivity, targeting physicians and the healthcare system while leaving broader epistemic trust intact. Rather than retreating from health information engagement, discrimination-exposed adults show elevated use of digital health channels, consistent with compensatory reorientation toward non-clinical information sources. These patients also experience meaningful deficits in health self-efficacy independent of their trust levels. Taken together, these findings describe engaged but institutionally alienated patients: motivated to manage their health yet deprived of the clinical trust that optimal care engagement requires. Restoring that trust will require sustained institutional accountability and structural reform, alongside digital health ecosystems designed to serve the patients who need them most.

## Data Availability

No data were generated by this study. All data underlying the findings are third-party data from existing public sources: the HINTS 6 (2022) and HINTS 7 (2024) public-use datasets, collected by the National Cancer Institute and freely available without restriction at https://hints.cancer.gov/data/download-data.aspx. The authors did not have any special access privileges that other researchers would not have. Analysis code is available from the corresponding author on reasonable request.

## Supporting information

**S1 File. Full regression results**. Complete coefficients, jackknife standard errors, 95% confidence intervals, and p-values for every model underlying Tables 2–5.

**S2 File. STROBE checklist**. Checklist of items for cross-sectional studies, with the location of each item in the manuscript.

## Acknowledgments

The authors thank the National Cancer Institute for collecting and providing public access to the HINTS data.

